# Peptide microarray based analysis of antibody responses to SARS-CoV-2 identifies unique epitopes with potential for diagnostic test development

**DOI:** 10.1101/2020.11.24.20216663

**Authors:** Pavlo Holenya, Paul Joris Lange, Ulf Reimer, Wolfram Woltersdorf, Thomas Panterodt, Michael Glas, Mark Wasner, Maren Eckey, Michael Drosch, Jörg-Michael Hollidt, Michael Naumann, Florian Kern, Holger Wenschuh, Robert Lange, Karsten Schnatbaum, Frank F. Bier

## Abstract

Humoral immunity to the Severe Adult Respiratory Syndrome (SARS) Coronavirus (CoV)-2 is not fully understood yet but may be a crucial factor of immune protection. The possibility of antibody cross-reactivity between SARS-CoV-2 and other human coronaviruses (HCoVs) would have important implications for immune protection but also for the development of specific diagnostic ELISA tests. Using peptide microarrays, n=24 patient samples and n=12 control samples were screened for antibodies against the entire SARS-CoV-2 proteome as well as the Spike (S), Nucleocapsid (N), VME1 (V), R1ab, and Protein 3a (AP3A) of the HCoV strains SARS, MERS, OC43 and 229E. While widespread cross-reactivity was revealed across several immunodominant regions of S and N, IgG binding to several SARS-CoV-2-derived peptides provided statistically significant discrimination between COVID-19 patients and controls. Selected target peptides may serve as capture antigens for future, highly COVID-19-specific diagnostic antibody tests.

## Introduction

The COVID-19 pandemic hit the world unprepared. At the time of writing global cases were in the millions and still rising [WHO, 2020]. Apart from social distancing, rapid testing of suspected cases and contacts is the key measure helping to contain the infection. The experience from many countries has shown that imprecise or incomplete knowledge of infection prevalence and viral transmission rates may result in a wrong assessment of the epidemiological situation, wrong predictions, and inadequate guidance and action with potentially grave consequences for societies and national economies. These observations underline the global need for reliable and highly specific tests.

SARS-CoV-2 infection is generally confirmed by RT-PCR on swabs taken from the nose and throat, however, virus load quickly becomes unmeasurable following the acute infection. Reliable serological monitoring will therefore be instrumental in determining the SARS-CoV-2 infection status of larger populations, determine existing immunity, and track local outbreaks [Pollán et al., 2020]. In the near future it may also greatly contribute to assessing vaccine-induced immunity at the population level. However, SARS-CoV-2 antibody testing is much more complex than appears at first glance. Currently available antibody tests are not very specific and may produce conflicting results [Krammer and Simon, 2020; Lisboa Bastos et al., 2020; Bond et al., 2020]. A plausible reason for that might be that antibody tests have generally been validated by samples from clinically symptomatic rather than mild or even asymptomatic cases. However, the reason for the overall unreliable performance of antibody tests is not fully understood. Interestingly, it has been suggested that, particularly in mild and asymptomatic cases, IgG responses may not be detectable until 3-4 weeks after infection [Fafi-Kremer et al., 2020] which may compound the issue.-

A large number of studies have examined antibody responses to SARS-CoV-2 [Siracusano et al., 2020]. Most of these used full-length proteins as capture antigens and thus have not identified reactivity at the single epitope level. Only few references [Amrun et al., 2020; Farrera-Soler et al. 2020; Poh et al., 2020; Wang et al., 2020; Zhang1 et al., 2020] including preprints [Zhang2 et al., 2020; Smith et al., 2020; Dahlke et al., 2020; Ladner et al., 2020] provide more detailed information on antibody specificity. Most of these studies comprise small sample numbers (n ≤ 10) and only one of them [Ladner et al., 2020] examined cross-reactivity with other HCoVs despite *in silico* analysis indicating that significant cross-reactivity is to be expected [Ahmed et al., 2020; Grifoni et al., 2020].

Here we present a first study of the humoral immune response against SARS-CoV-2 at the single linear peptide epitope level covering all SARS-CoV-2 antigens as well as the S, N, E and M proteins of the pandemic/epidemic-related human Severe Adult Respiratory Syndrome and Middle East Respiratory Syndrome-related human coronaviruses (SARS-CoV and MERS-CoV), and, in addition, the seasonal cold-related strains human coronavirus OC43 and 229E. Our results provide deeper insight into the immune response to HCoVs than previously published work which could pave the way to SARS-CoV-2 specific and reliable COVID-19 diagnostic test development.

## Results

### Identification of immunodominant epitopes

Serum samples from COVID-19 patients analyzed by peptide microarray technology exhibited high levels of antibody reactivity, whereas almost no reactivity was detected with control samples. Several immunodominant regions were identified. These were located primarily on the SARS2 S and N proteins (**Figure 1, Supplementary Figures 1-6**). There was also strong IgG reactivity to several peptides derived from the SARS2 M, AP3A and R1AB proteins, however, immune dominance was less pronounced in these proteins (**Supplementary Figures 3, 5 and 6)**. All remaining SARS2 antigens demonstrated no or only minor immunogenicity in terms of IgG response.

**Figure 1.**
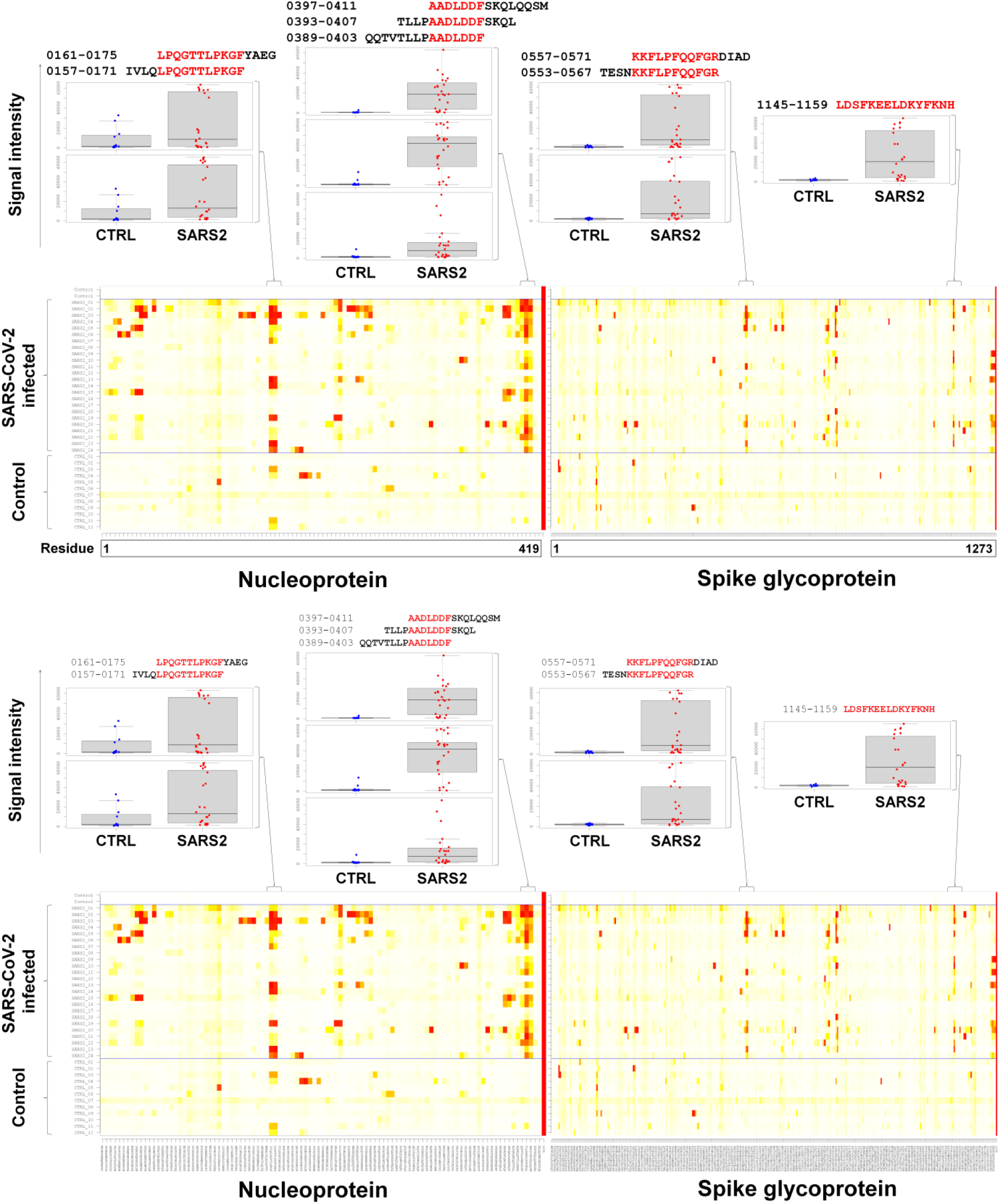
Peptide microarray based analysis of IgG responses to SARS-CoV-2 Nucleoprotein (N) and Spike glycoprotein (S) measured in human sera. Heatmaps represent two groups of serum samples: SARS-CoV-2 infected and control groups. The two upper rows of the heatmaps reflect signals obtained with detection antibody only (no serum). Columns represent peptide sequences, rows represent samples. Colours indicate the signal values obtained from triplicate spots ranging from white (0 or low intensity) over yellow (middle intensity) to red (maximal intensity of 65535 light units). The two upper rows of the heatmaps reflect signals obtained with detection antibody only (no serum). The right column in each heatmap reflects the highest possible signal intensity. Box plots show the signal distribution in each group for selected immunodominant regions.

For quantitative characterization of the identified immunodominant regions, responses in SARS-CoV-2 infected individuals were compared to those in the control group for each single peptide and for combinations of three, four and five peptides by the non-parametric Wilcoxon Rank Sum test. Results for selected peptide hits and the best combinations thereof are presented in **Figure 2. Table 1** shows a selection of identified epitopes represented either by single peptides or by common core sequences derived from two or more overlapping peptides. To narrow down the identified immunodominant peptides to the potential immunodominant epitope sequences, the peptides with the highest accuracy scores (calculated as the sum of true positive and true negative divided by the total number of observations) were listed in the order of appearance in the corresponding antigens. If applicable, sequence overlaps were marked in red. A complete list of peptides showing the most significant discrimination between SARS-CoV-2 infected and the control group can be found in **Supplementary Table 2**.

**Table 1.**
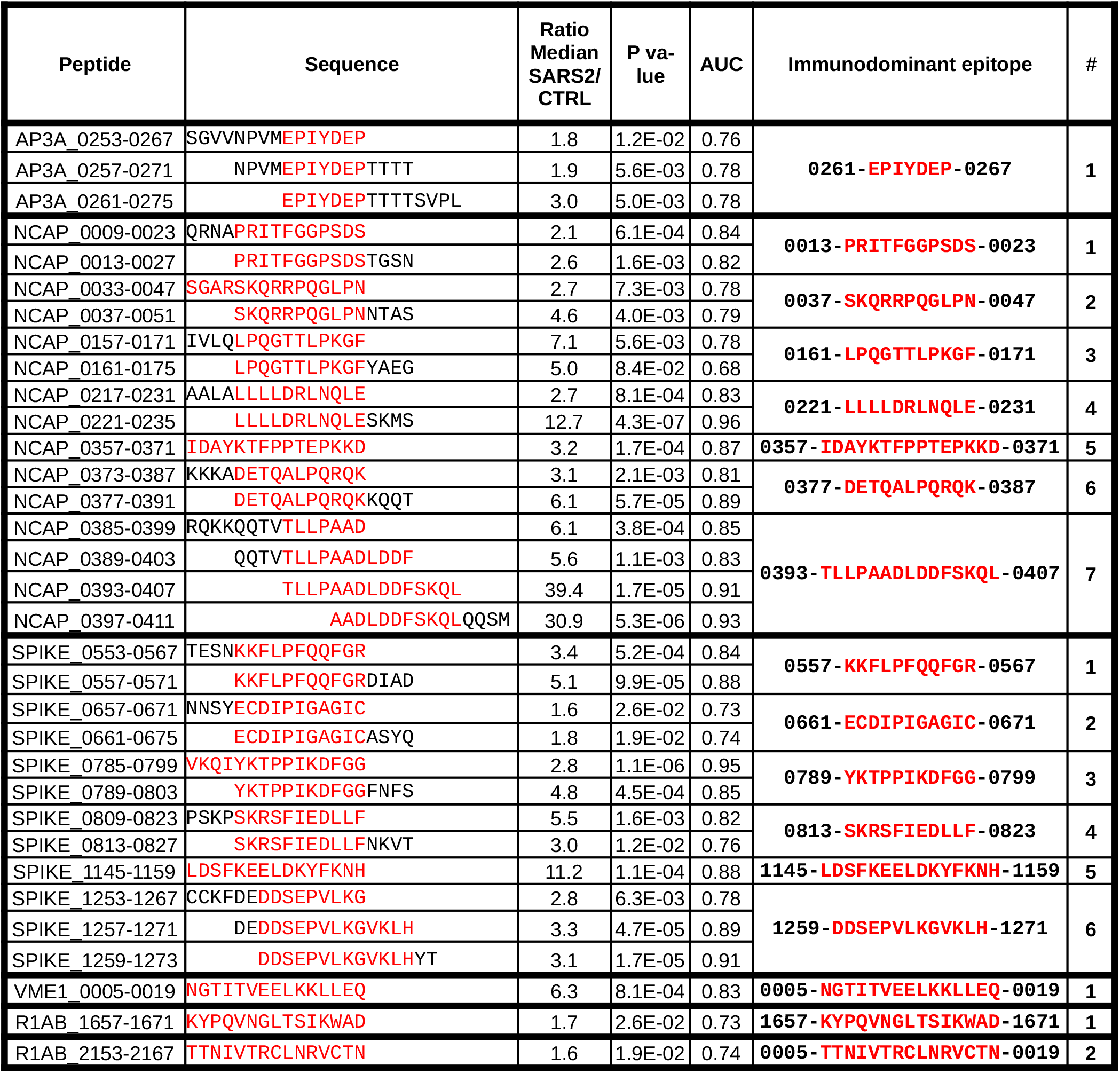
Selected immunodominant SARS-CoV-2 epitopes identified by peptide microarrays. Selected peptides providing good separation between the groups are listed. Immunodominant epitopes (highlighted red) are derived from the sequences of (overlapping) peptides.

**Figure 2.**
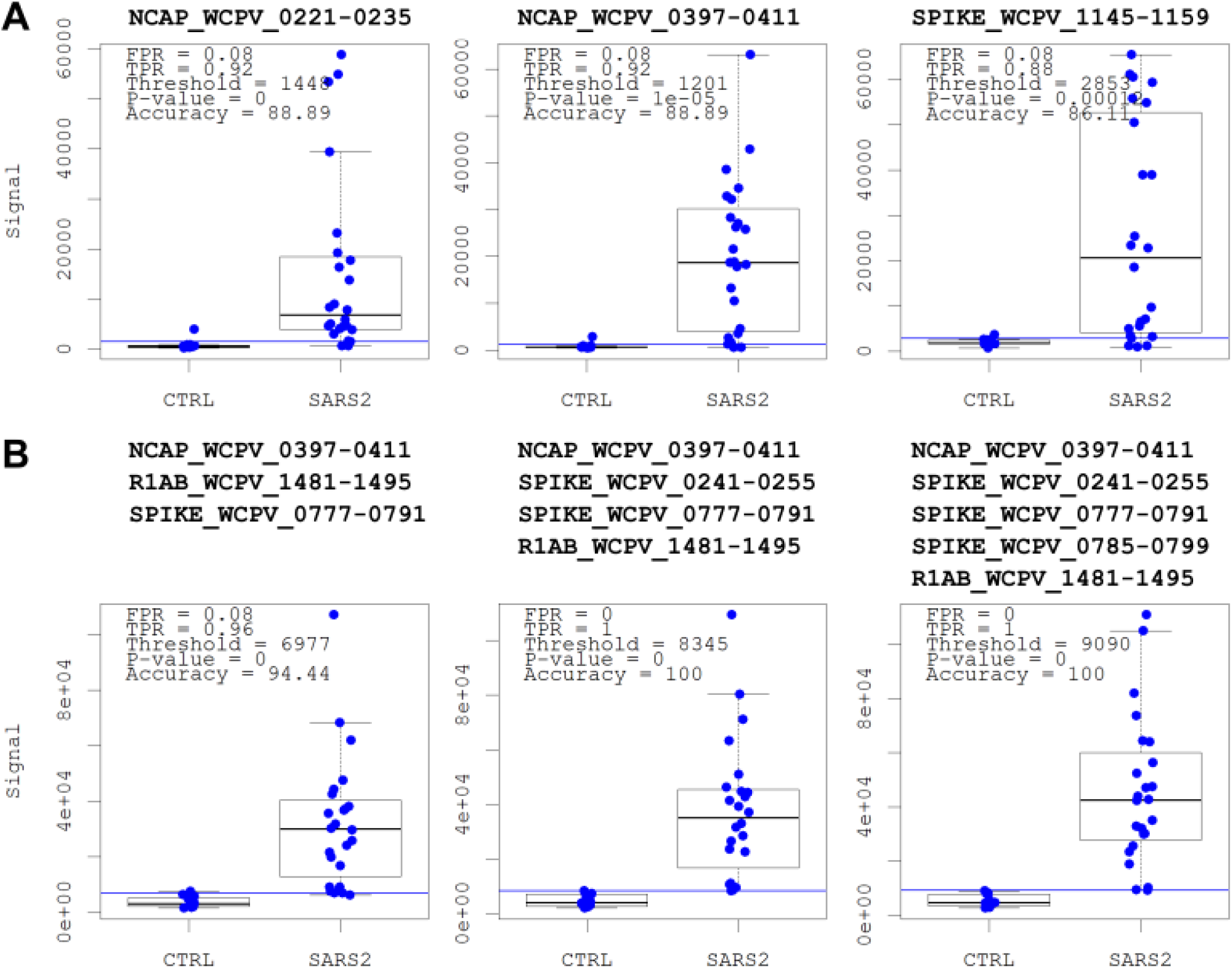
Selected single peptides and combination of peptides providing the best discrimination between sample groups. A. For each peptide the signal intensity was compared between the SARS-CoV2 infected (“SARS2”) and the control (CTRL) group by Wilcoxon Rank Sum test with the box plots showing the ability of selected peptides to discriminate between groups. B. Combinations of three, four and five peptides providing the best discrimination between the groups by Wilcoxon Rank Sum test. Each data point represents the sum of the signals of the combined peptides.

### Microarray assay results strongly correlate with a commonly used diagnostic test

To evaluate the diagnostic potential of a peptide-based assay, a comparison with a commonly used diagnostic test utilizing a full-length viral antigen was performed. **Figure 3** shows the quantitative results obtained for the SARS-CoV-2 infected group with a) the commercial ELISA test (EUROIMMUN) and b) the peptide microarray. Both assays demonstrated a strong correlation with a Spearman’s rank correlation coefficient of 0.88.

**Figure 3.**
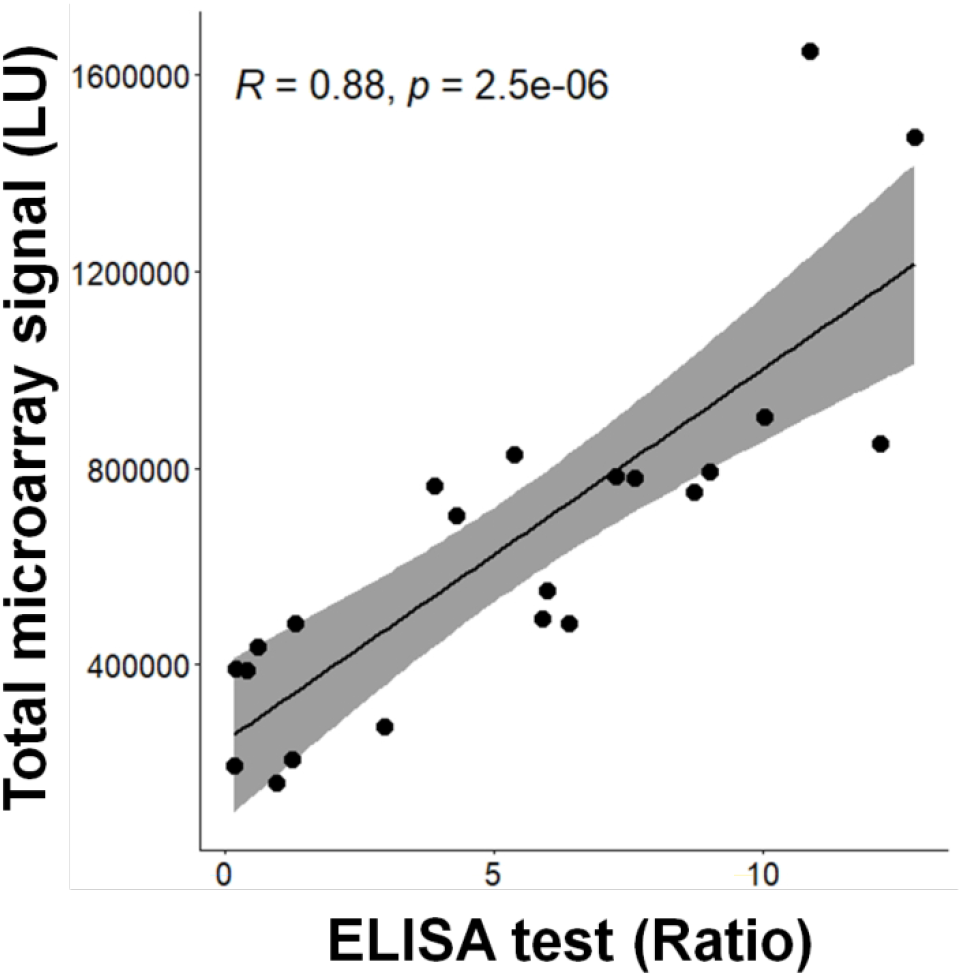
Correlation between the commercial ELISA and the peptide microarray signals. The Spearman’s correlation coefficient between the ELISA signal and the summed microarray signals for SARS-CoV-2 S and N peptides was calculated for sera from SARS-CoV-2 infected patients. In the ELISA assay, which used a recombinant S glycoprotein as capture antigen, the quotient of the extinction of the patient sample in comparison to the calibrator was used as net assay signal. The total microarray signals for each sample were calculated as a sum of all corresponding signal intensities above the upper 10^−16^% quantile of the noise distribution.

### Cross-reactivity with seasonal common cold coronaviruses experimentally confirmed

Antibody cross-reactivity between SARS-CoV-2 and seasonal common cold coronaviruses may be highly important with respect to the clinical course of COVID-19 and at the same time represents a challenge for the development of a specific test. We, therefore, analyzed IgG reactivity to peptides derived from the S, N, E, and M proteins of SARS-CoV, MERS-CoV, HCoV-OC43 and HCoV-229E (**Supplementary Table 1**) in both groups. This revealed highly similar IgG binding profiles against the N and M proteins of SARS-CoV-2 and SARS-CoV, whereas the corresponding proteins of the remaining viral strains showed only weak and scattered signals (**Supplementary Figures 1 and 3**). Most notably, peptides derived from the S protein of all viral strains showed clear signal patterns among SARS-CoV-2 positive patient samples, suggesting that this antigen contained cross-reactive determinants (**Supplementary Figure 2**). IgG reactivity was directed at two sequence regions in particular (**Figure 4**). Each of these regions was represented by overlapping peptides containing highly conserved cross-reactive epitopes (**Figure 4, right panel**). One consensus motif, **R**_**0815**_**S-IED-LF**_**0823**_ (numbers referring to the location of the epitope in the SARS2 antigen) was present in all five viruses, and a second consensus motif, **F**_**1148**_**--ELD--FKN**_**1158**_ was found in the viruses SARS-CoV-2, SARS-CoV, MERS-Cov and HCoV-OC43, but not HCoV-229E.

**Figure 4.**
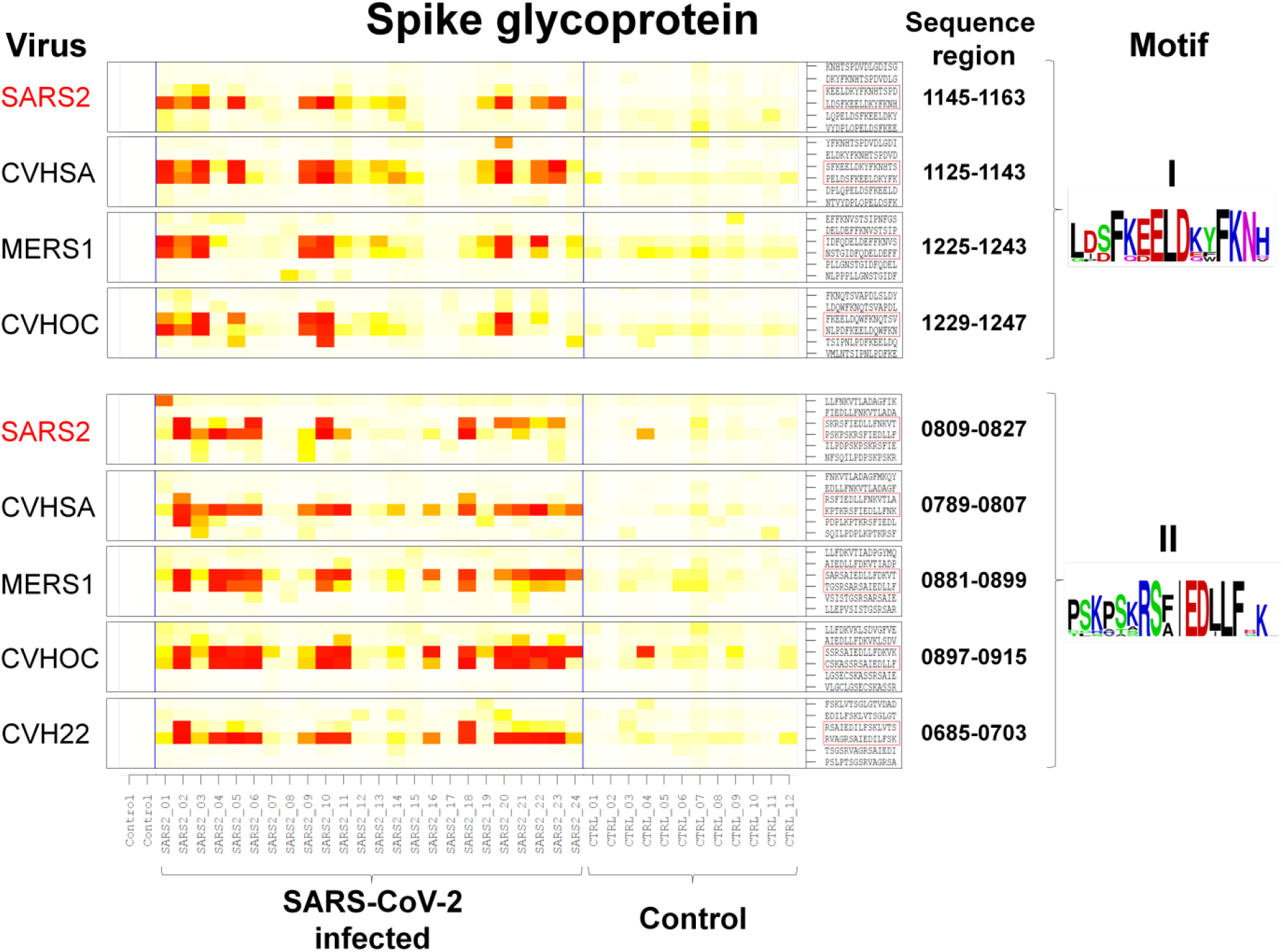
Serologic cross-reactivity between SARS-CoV-2 and other Coronaviruses. Left: Conserved S protein regions of different coronaviruses showing a strong IgG seroreactivity in sera from SARS-CoV-2 infected patients. Each region is represented by two overlapping peptides (framed red). Right: cross-reactive motifs resulting from sequence alignment of the peptides re-flecting a high homology. Motif II was present in all five viruses, whereas motif I was found in the viruses SARS-CoV-2, SARS-CoV, MERS-Cov and HCoV-OC43, but not HCoV-229E.

To further analyze serologic cross-reactivity of SARS-CoV-2 positive patients against common cold coronaviruses we performed a gapless alignment of the N and S sequences from all coronaviruses (**Figure 5**). For each strain, this yielded the sequence regions that were complementary to the previously identified SARS-CoV-2 epitopes. Finally, for each of the amino acid residues of the thus identified sequences, the median signal of all overlap - ping peptides containing that residue was calculated and visualized by the same colour coding as in the previously presented heatmaps (**Figure 5**). The immunodominant epitopes #6 and #7 of the N protein and #1 and #3 of the S protein of SARS-CoV-2 (see **Table 1** for a list of the sequences) displayed strong signals with samples from SARS-CoV-2 infected individuals compared to the control samples. At the same time, no signals were found for the homologous sequences of the other coronavirus strains, indicating that IgG responses to these epitopes may be truly specific for SARS-CoV-2 (**Figure 5**, Top). SARS-CoV-2 epitopes #1-#5 of the N and #2 and #6 of the S protein showed a strong homology to SARS-CoV which resulted in serologic cross-reactivity to this virus (**Figure 5**, Middle). Finally, epitopes #4 and #5 of the S protein were the least SARS-CoV-2 specific and exhibited considerable cross-reactivity between all sequence variants (**Figure 5**, Bottom).

**Figure 5.**
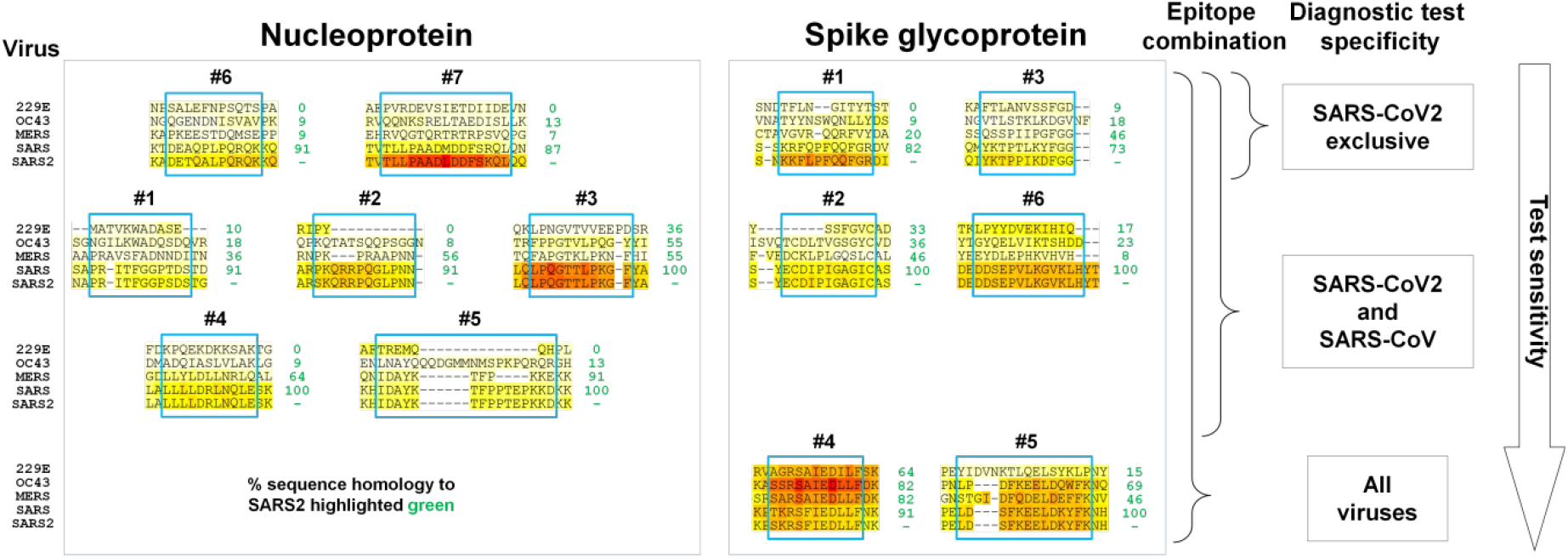
Potential Nucleoprotein (N) and Spike glycoprotein (S) epitope combinations for a diagnostic test. Each epitope (depicted with the same numbering as in Figure 2) is represented by a heatmap showing aligned sequences of the investigated coronaviruses. Blue frames indicate the inferred epitope length. The colored background of each amino acid residue reflects the median signal intensity of the SARS-CoV-2 positive cohort calculated from signals of all peptides on the microarray which contained this residue. The color code ranges from white (0 or low intensity) over yellow (middle intensity) to red (maximal intensity of 65535 light units). Per cent sequence homology to SARS-CoV-2 is indicated in green. Based on the observed cross-reactivity of the SARS-CoV-2-induced IgG responses toward the other virus types, certain combinations of SARS-CoV-2 epitopes may be used to develop diagnostic tests with different specificities.

## Discussion

The work presented here used a systematic approach that provides deeper insight into the humoral immune response to HCoVs and lays a fundament for the development of specific serological tests of SARS-CoV-2 infection. The approach is based on four pillars: (i) peptide microarray based analysis of the humoral immune response against the entire SARS-CoV-2 proteome and other HCoV proteomes, (ii) identification of all immunodominant epitopes at high resolution, (iii) selection of SARS-CoV-2 peptides that are recognized by serum antibodies of COVID-19 patients but not by serum antibodies from controls and (iv) selection of SARS-CoV-2 peptides whose homologous peptides in other HCoVs are not recognized by serum antibodies from COVID-19 patients.

Our analysis of IgG responses of COVID-19 patients against all peptides of the SARS-CoV-2 proteome revealed that IgG binding was mainly focused on both the S and N proteins, indicating that these two proteins are indeed immunodominant (**Figure 1, Supplementary Figures 1-6**). The fact that the M, AP3A and R1ab proteins were also recognized, however, shows that responses are not exclusive to S and N. It remains to be resolved in future studies if the most dominant immune responses against certain antigens are also the most effective responses concerning disease protection.

Applying stringent bioinformatic criteria we identified several linear epitopes that were immunodominant among SARS-CoV-2 positive patients (**Table 1**). Previous analyses of antibody epitopes in SARS-CoV revealed that many such epitopes were indeed linear [Guo et al., 2004; Shichijo et al., 2004]. Interestingly, some immunodominant regions in the N and S proteins that have been previously reported in the literature [Poh et al., 2020; Amrun et al., 2020] correspond closely to epitopes #3 from N and #1 and #4 from S (**Table 1**), respectively. This confirms our findings and illustrates the potential of the peptide microarrays for epitope identification. Longer sequence stretches encompassing some of the epitopes identified here have also been described in published manuscripts [Wang et al., 2020; Farrera-Soler et al., 2020; Zhang1 et al., 2020] or in non-peer-reviewed preprint publications, however, without narrowing them down [Zhang2 et al., 2020; Smith et al., 2020; Dahlke et al., 2020]. With our comprehensive peptide library we further completed the list of the previously described immunodominant SARS-CoV-2 regions and specified the corresponding epitopes.

The most important question with respect to developing a reliable diagnostic test was, however, whether the IgG response to the identified SARS2-specific peptides can effectively distinguish COVID-19 patients and controls. This was indeed the case, as a comparison of serum reactivity in COVID-19 patients versus the control group, peptide by peptide, identified many peptides to which responses were significantly different between the groups (**Table 1, Figure 2A**). The combination of selected peptides derived from different SARS-CoV-2 antigens further improved the discrimination between the sample groups as shown in **Figure 2B**, and also might increase sensitivity when applied in a diagnostic test. As a further step to assess the diagnostic potential of the obtained data, the quantitative results obtained with the peptide microarrays were compared with those generated by a commercial ELISA test (**Figure 3**). We observed a very strong correlation what suggests that a combination of short linear synthetic peptides may be used for the development of a diagnostic test instead of full-length antigens. Moreover, as it was found that only a small proportion of all peptides forms B cell epitopes, the selection of peptides must not necessarily span the whole antigen, but might be limited to immunodominant regions identified by a peptide microarray screening.

Multiple studies utilizing *in silico* prediction of SARS-CoV-2 immunogenicity [Wang et al., 2020; Yuan et al., 2020; Palm et al., 2020; Smith et al., 2020] postulated the existence of cross-reactive immunity between different coronaviruses, in particular between SARS-CoV-2 and SARS-CoV, because several known B-cell and T-cell target peptides in SARS-CoV exist almost identically in SARS-CoV-2. Cross-reactive T cell epitopes were experimentally confirmed by several authors [Braun et al., 2020; Mateus et al., 2020] and some degree of B cell cross reactivity to HCoVs has been identified by western blot analysis of full length antigens [Bonifacius et al., 2020]. However, there is still a lack of experimental data identifying cross-reactive B cell epitopes. We found two immunodominant regions in the SARS-CoV-2 S protein (1148-FKEELDKYFKN-1158 and 0815-RSFIEDLLF-0823) which were highly cross-reactive with the prevalent seasonal “common cold” coronaviruses HCoV-OC43 and HCoV-229E (**Figure 4**). A similar result obtained with a different assay based on 30-mer peptide cDNA-fusion pools in a non-peer review publication confirms our findings [Ladner et al., 2020].

The identified cross-reactivity is of particular interest, as it suggests that exposure to common cold HCoVs may provide a degree of immunity to SARS-CoV-2. The presence of such preexisting immunity may have a positive impact on the course and outcome of COVID-19. Arguments against this hypothesis are, however, that it has been reported that the common cold coronaviruses generate only a weak and transient humoral immune response [Huang et al., 2020] and that reconvalescents from SARS1 – a virus that is structurally more closely related to SARS2 than the common cold coronaviruses – did not have SARS2 neutralization titers in plasma [Lv et al., 2020]. Of note, preexisting immunity could also be detrimental, as it may inhibit production of antibodies with certain specificity, *i*.*e*. a phenomenon called original antigenic sin. This phenomenon may be responsible for ineffective vaccine response to a recently developed Cytomegalovirus vaccine [Baraniak et al., 2019]. Even worse, existing antibodies could promote infection of target cells by facilitating viral uptake, a phenomenon called antibody-dependent enhancement [Fierz et al., 2020; Arvin et al., 2020]. Our results demonstrate that some of the patient sera, *e*.*g*. numbers SARS_20 to SARS_23, show stronger reactivity towards the HCoV-OC43 variant of the cross-reactive epitope II than to the respective SARS-CoV-2 epitope (**Figure 4**). This is an interesting observation as there is no other logical explanation for this than that the antibody was made in response to the HCoV-OC43 virus in a previous HCoV-OC43 infection. A number of additional peptides that led to strong IgG responses (**Figure 4**) support the notion of preexisting B cell immunity to SARS-CoV-2 in those exposed to other coronaviruses. It will be up to future studies with larger cohorts to determine whether preexisting immunity has an effect on disease severity.

While the physiological effects of serologic cross-reactivity require further studies, it is obvious that it makes the development of reliable antibody tests more difficult. Antibody based ELISA tests that use full-length conformational antigens or parts thereof as capture antigens might give false positive results in individuals with previous exposure to common cold coronaviruses. In this regard, the usage of the S protein antigen in a diagnostic test might be particularly problematic due to several known homologous regions between SARS-CoV-2 and other coronaviruses. The detailed analysis of IgG reactivity against SARS-CoV, MERS-CoV, HCoV-OC43 and HCoV-229E S and N proteins indeed revealed widespread cross-reactivity in several immunodominant regions (**Figure 4**), which is most probably a consequence of sequence homology. However, the observed cross-reactivity could not be predicted by simple sequence alignment unless 100% homology was present. With a view to develop a diagnostic test, our finding that epitopes #6 and #7 of N and #1 and #3 of S appear to provide a high specificity for SARS-CoV-2 (**Figure 5**) is of major interest. A combination of such epitopes in a peptide ELISA may increase sensitivity without increasing the risk of false positives.

In conclusion, our study experimentally confirms antibody cross-reactivity between SARS-CoV-2 and other HCoVs, but at the same time reveals antibody reactivity to several epitopes that are unique to SARS-CoV-2. These epitopes are of particular interest for the development of diagnostic tests. Additional, larger studies are required to confirm these results and accelerate the development of a sensitive and specific test for SARS-CoV-2 infection that we urgently need in order to deal with the current and possible future waves of this pandemic.

## Data Availability

all data are presented in the manuscript

## Acknowledgements

The authors are grateful for the assistance of the following local health authorities (Gesundheitsämter) of Landkreis Börde (Frau Dr. Kontzog and Team), Landkreis Salz-landkreis (Frau Leonhardt and Team), and Landkreis Harz (Frau Dr. Christiansen and Team) for their support in identifying appropriate blood donors.

## Author Contributions

W.W., T.P., M.G., M.W. collected samples; P.H., P.J.L. conducted the experiments; P.H., P.J.L.; U.R. performed data analysis; M.N., R.L., F.B. initiated and projected the study; P.H., U.R., M.E., M.D., J.M.H, M.N., F.K., H.W., R.L, K.S., F.B. designed the experiments; P.H., P.J.L., F.K., R.L., K.S., F.B. wrote the paper.

## METHOD DETAILS

### Blood sampling

Following written informed consent peripheral blood samples were obtained by vene puncture / from periperal venous catheters. Multiple samples were collected into a Vacutainer Tube (Serum-separating tube) with a volume of 8 ml. Serum was collected after centrifugation, samples were allowed to clot for 1 hour, and the clot was removed by centrifugation. The samples were divided and stored in aliquots at −80°C until use.

### RT-PCR RNA collection according to Guidelines of the CDC

Samples were collected from the upper respiratory tract using a nasopharyngeal (NP) with plastic by trained medical staff. In the case of the NP the swab was inserted deeply into the nostril until resistance was encountered and the depth was equal to the distance from the nose to the ears of the patients. The swab was gently rubbed against the nasopharynx and left there in order to absorb enough secret. Swabs were then immediately placed into a sterile transport tube. Samples were cooled and tested no more than 48 hours after collection.

### Participants

Controls: Controls consist of excess serum samples collected in accordance to the regulations for the use of rest samples for scientific purposes at the KH Labor. All samples have been collected from form patients hospitalized in April 2020. Sera in the control group tested negative for COVID-19 infection in the EUROIMMUN IgG Elisa Test. We used samples from different specialties, including cardiology, gastroenterology, nephrology and the intensive care unit.

SARS-CoV-2 infected group: COVID-19 patients (n=13 female, age range 23 to 70 years; n=11 male, age range 35 to 70 years) were recruited across the spectrum of clinical severity, ranging from asymptomatic/mild to severe/SARS. COVID-19 diagnosis was confirmed either by diagnostic PCR and/or commercial ELISA carried out 14 days or later after the onset of symptoms. Detailed information on all patients and samples is provided in **Supplementary Table 3**.

Inclusion/exclusion criteria were: Fully conversant in the German or English language, able and willing to attend to the study requirements, written informed consent to participate in this study / history of four or more infections with antibiotic treatment in the last year, severe liver or kidney disease, patients with immunotherapy, patients that suffer from any oncological disease.

To ensure valid test results we screened patients according to the specified inclusion and exclusion criteria. All Patients were interviewed systematically according to an anamnestic questionnaire. To validate COVID-19 cases, RT-PCR, clinical diagnosis and the result of the Euroimmune IgG Assay were used in conjunction. Patients were questioned about the time of onset of symptoms. All Patients included in this study could precisely specify at which date symptoms occurred. After the onset of symptoms, all but one Patient underwent RT-PCR testing and tested positive for COVID-19. We ensured that no patient took medicine or suffered from a disease that could potentially affect the development of antibodies. The severity of COVID-19 was determined on the basis of WHO guidelines into asymptomatic, mild (common cold-like symptoms), moderate (dyspnea, clinical signs of pulmonary distress) and severe (requiring oxygen supplementation, intensive care with or without ventilation) [WHO Global, 2020].

### Peptide microarrays

Antibody response profiling was performed with a peptide microarray (JPT Peptide Technologies GmbH) containing 5513 peptides covering the full proteome of SARS-CoV-2 and, in addition, the S, N, E, and M proteins of the following HCoVs: SARS-CoV-1, MERS-CoV, HCov 229E, OC43. All proteins were represented by overlapping peptides covering the entire protein sequence (15 aa length, 11 aa overlap). All proteins covered by the peptide library are listed in **Supplementary Table 1**.

### Peptide synthesis and microarray production

Peptides were synthesized and immobilized on peptide microarray slides as described previously [Stephenson et al., 2015]. In brief, the peptides were synthesized using SPOT synthesis [Wenschuh et al., 2000], cleaved from the solid support and chemoselectively immobilized on functionalized glass slides. Each peptide was deposited on the microarray in triplicates.

### Microarray assay and data analysis

The peptide microarrays were incubated with sera (applied dilution 1:200) in an HS 4800 microarray processing station (Tecan) for two hours at 30°C, followed by incubation with 0.1 μg/mL fluorescently labelled anti human IgG deg/mL fluorescently labelled anti human IgG detection antibody (Jackson Immunoresearch, 109-605-098). Washing steps were performed prior to every incubation step with 0.1% Tween-20 in 1x TBS. After the final incubation step the microarrays were washed (0.05% Tween-20 in 0.1x SSC) and dried in a stream of nitrogen. Each microarray was scanned using a GenePix Autoloader 4300 SL50 (Molecular Devices, Pixel size: 10 μg/mL fluorescently labelled anti human IgG dem). Signal intensities were evaluated using GenePix Pro 7.0 analysis software (Molecular Devices). For each peptide, the MMC2 value of the three triplicates was calculated. The MMC2 value was equal to the mean value of all three instances on the microarray except when the coefficient of variation (CV) – standard-deviation divided by the mean value – was larger than 0.5. In this case the mean of the two values closest to each other (MC2) was assigned to MMC2. Further data analysis and generation of the heatmapts was performed using the statistical computing and graphics software R (Version 4.0.2, www.r-project.org).

### RT-PCR test

We used the protocol described by the centers for disease control and prevention for the CDC 2019 Novel Coronavirus (2019-nCoV) Real-Time Reverse Transcriptase (RT)–PCR Diagnostic Panel.

### ELISA test

To detect antibodies, we used the Euroimmun Anti-SARS-CoV-2 ELISA (IgG), which was manufactured by Euroimmun Medizinische Labordiagnostika AG, Lübeck, Germany. The break-off microplate wells were covered with the antigen, a recombinant structural spike 1 (S1) protein of SARS-CoV-2 expressed in HEK293 [FDA, 2020]. First, the diluted patient samples were incubated in the wells which lead to specific IgG antibodies binding to the antigens. In order to detect the bound antibodies, an enzyme-labelled anti-human IgG antibody detected antigen-antibody complexes and catalysed a colour reaction. By calculating the extinction of the sample over the extinction of the calibrator the results could be evaluated semi-quantitatively. A ratio below 0.8 was considered a negative result. A ratio between 0.8 and 1.1 was considered a borderline result. A ratio above 1.1 was considered a positive result [Euroimmun Medizinische Labordiagnostika AG, 2020].

## QUANTIFICATION AND STATISTICAL ANALYSIS

Data analysis and heatmap generation was performed using the statistical computing and graphics software R (Version 4.0.2, www.r-project.org).

The data set used for statistical analysis of the microarray results and for generation of all heatmap presentations was based on MMC2 values calculated from signals of the triplicate spots of each individual peptide. The MMC2 value calculation is described in detail in the ***Microarray assay and data analysis*** section.

The principle of the colour coding of all heatmaps is described in the corresponding figure legends. The coloured fields in the heatmaps reflect the calculated MMC2 values of each individual peptide.

The boxplots in **Figure 1** and **Figure 2** show the median of the corresponding sample groups. The central rectangles span the first quartile to the third quartile (the *interquartile range* or *IQR*). The whiskers represent the following values: upper whisker = min(max(x), Quartile 3 + 1.5 * IQR); lower whisker = max(min(x), Quartile 1 – 1.5 * IQR). The points in the boxplots show the MMC2 values for the individual samples.

Statistical comparison of two sample groups was done by the Wilcoxon Rank Sum test (R package *stats*, n=24 patient samples and n=12 control samples). ROC analysis was performed using the R package *ROCR*. The selection of the peptide hits with significant separation between the sample groups was based on the accuracy value calculated as ‘(true positive + true negative)/total number of observations’. An accuracy value of ≥0.4 was considered to indicate immunogenicity (**Supplementary Table 2**). Immunodominance was assigned to epitopes based on the highest accuracy scores and the sequences of the corresponding overlapping peptides (**Table 1**).

The Spearman’s rank correlation coefficient reflecting the relationship between the commercial ELISA and the peptide microarray (**Figure 3**) was calculated as described in the figure legend using the R package *ggpubr*.

---

**Supplementary Figure 1.**
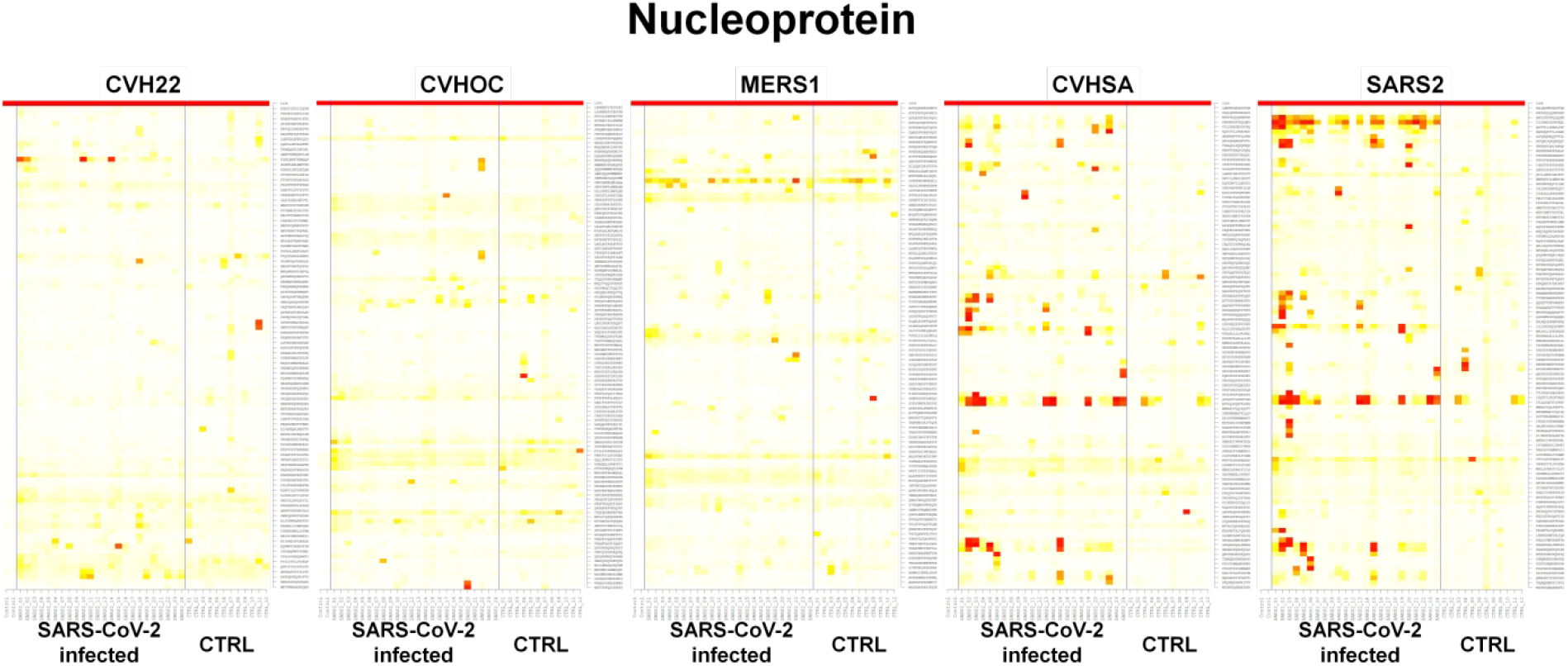
Peptide microarray analysis of IgG responses to Nucleoprotein (N) of different coronaviruses. Heatmaps represent two groups of serum samples: SARS-CoV-2 infected and control groups. The two left rows of the heatmaps reflect signals obtained with detection antibody only (no serum). Columns represent peptide sequences, rows represent samples. Colours indicate the signal values obtained from triplicate spots ranging from white (0 or low intensity) over yellow (middle intensity) to red (maximal intensity of 65535 light units). The two left rows of the heatmaps reflect signals obtained with detection antibody only (no serum). The upper row in each heatmap reflects the highest possible signal intensity.

**Supplementary Figure 2.**
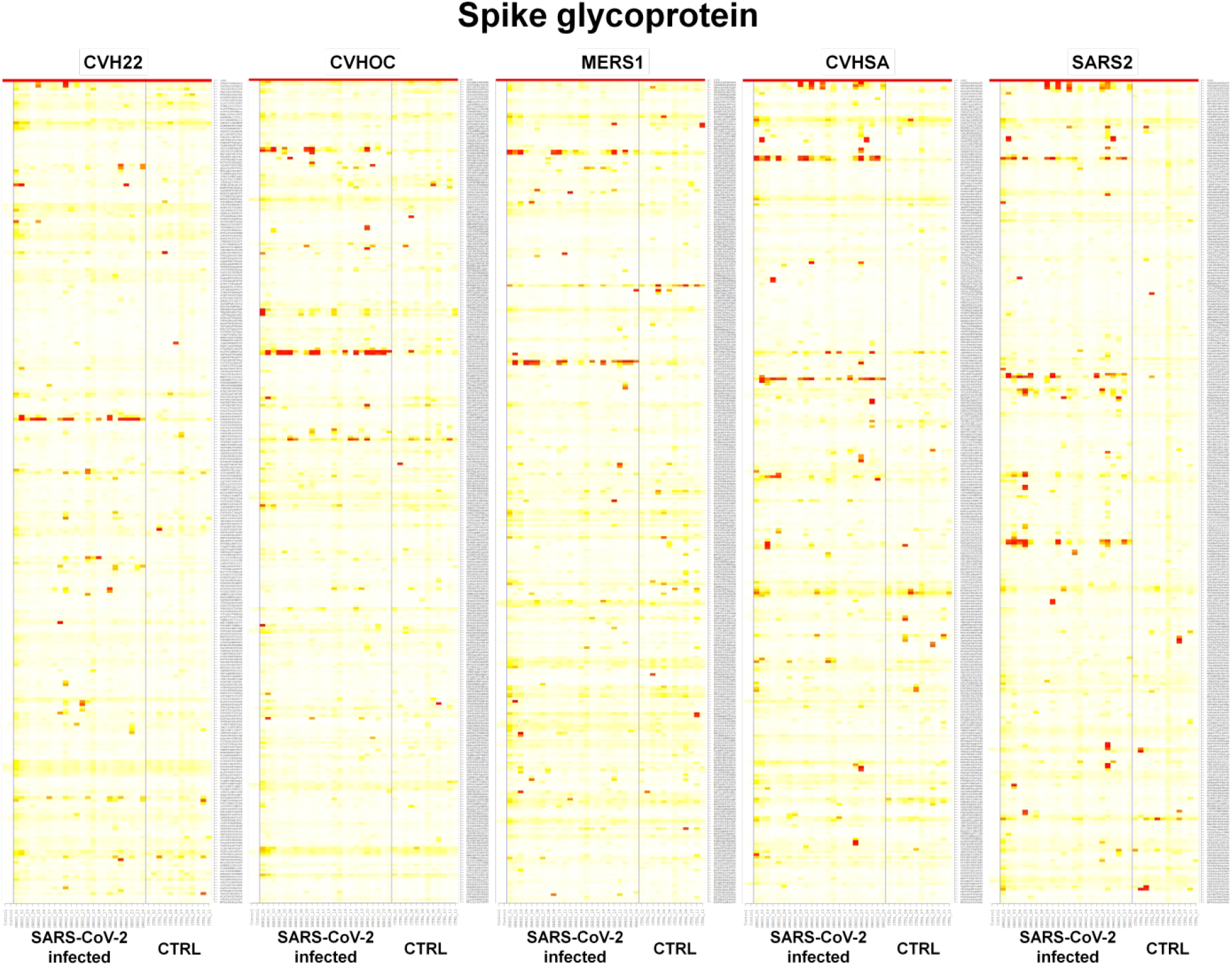
Peptide microarray analysis of IgG responses to Spike glycoprotein (S) of different coronaviruses. Heatmaps represent two groups of serum samples: SARS-CoV-2 infected and control groups. The two left rows of the heatmaps reflect signals obtained with detection antibody only (no serum). Columns represent peptide sequences, rows represent samples. Colours indicate the signal values obtained from triplicate spots ranging from white (0 or low intensity) over yellow (middle intensity) to red (maximal intensity of 65535 light units). The two left rows of the heatmaps reflect signals obtained with detection antibody only (no serum). The upper row in each heatmap reflects the highest possible signal intensity.

**Supplementary Figure 3.**
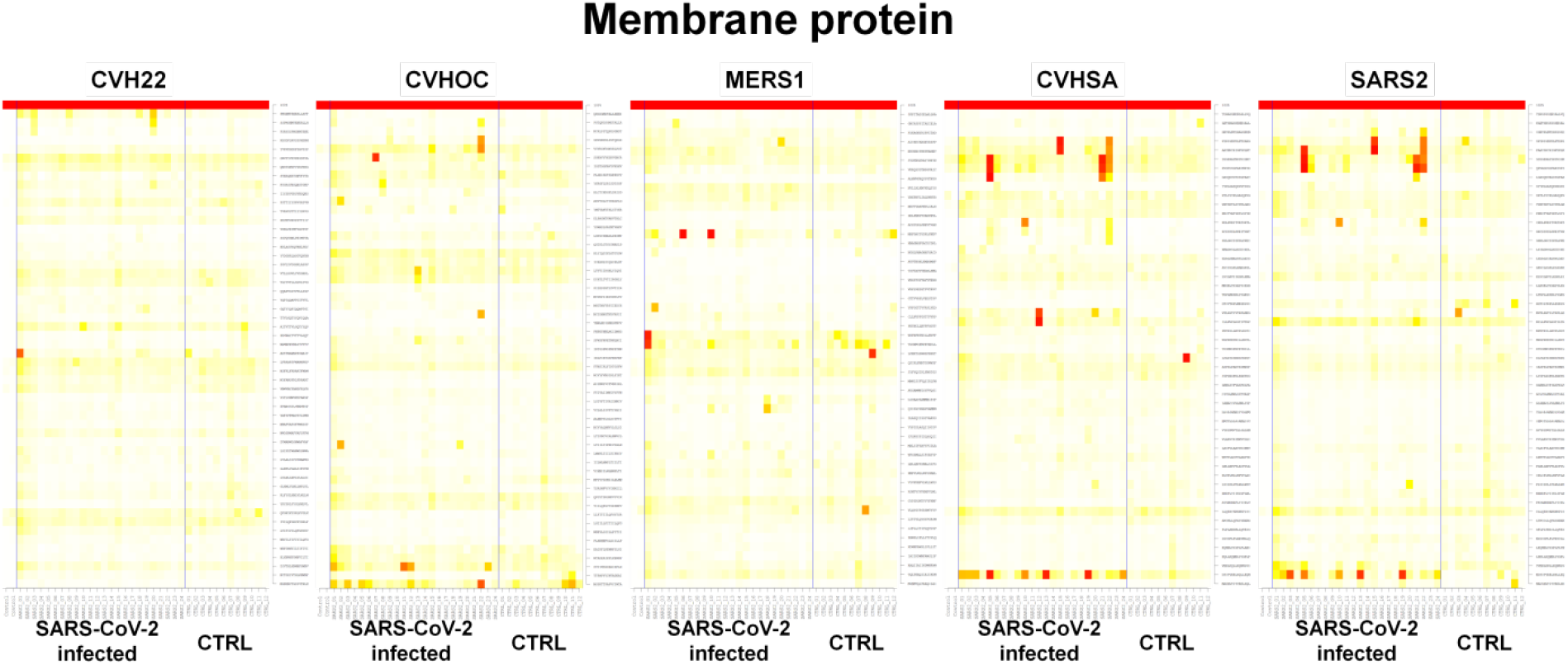
Peptide microarray analysis of IgG responses to Membrane protein (M) of different coronaviruses. Heatmaps represent two groups of serum samples: SARS-CoV-2 infected and control groups. The two left rows of the heatmaps reflect signals obtained with detection antibody only (no serum). Columns represent peptide sequences, rows represent samples. Colours indicate the signal values obtained from triplicate spots ranging from white (0 or low intensity) over yellow (middle intensity) to red (maximal intensity of 65535 light units). The two left rows of the heatmaps reflect signals obtained with detection antibody only (no serum). The upper row in each heatmap reflects the highest possible signal intensity.

**Supplementary Figure 4.**
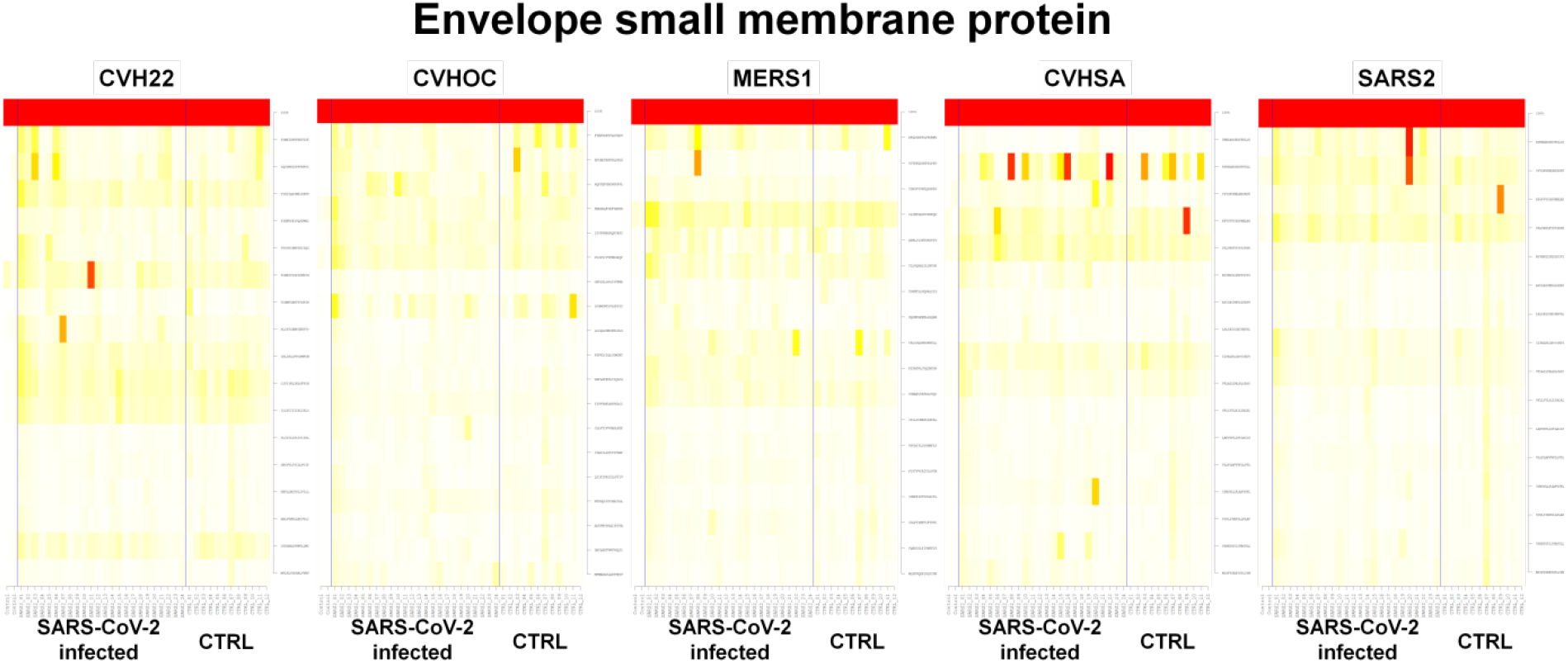
Peptide microarray analysis of IgG responses to envelope small membrane protein (E) of different coronaviruses. Heatmaps represent two groups of serum samples: SARS-CoV-2 infected and control groups. The two left rows of the heatmaps reflect signals obtained with detection antibody only (no serum). Columns represent peptide sequences, rows represent samples. Colours indicate the signal values obtained from triplicate spots ranging from white (0 or low intensity) over yellow (middle intensity) to red (maximal intensity of 65535 light units). The two left rows of the heatmaps reflect signals obtained with detection antibody only (no serum). The upper row in each heatmap reflects the highest possible signal intensity.

**Supplementary Figure 5.**
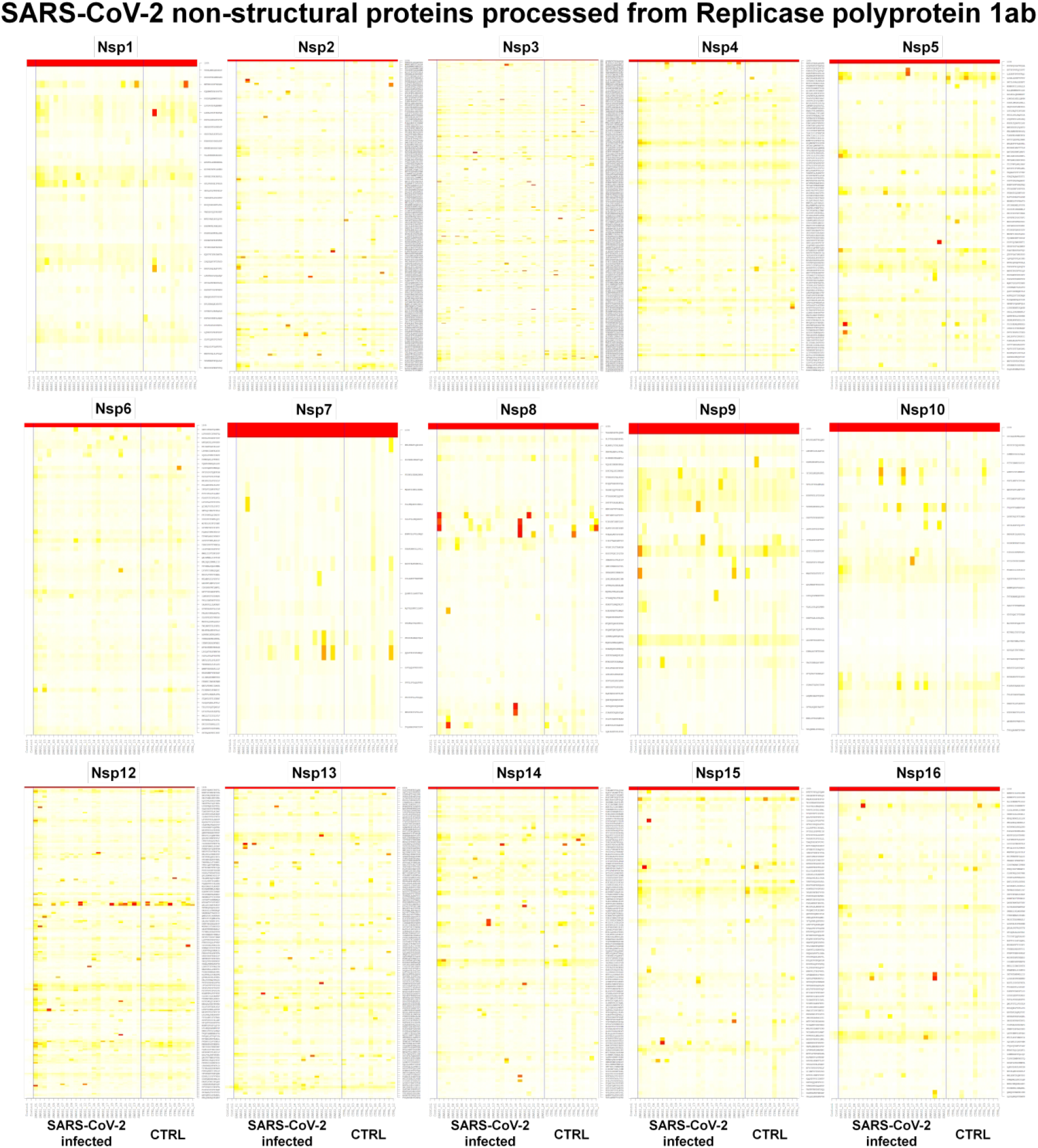
Peptide microarray analysis of IgG responses to SARS-CoV-2 non-structural proteins processed from Replicase polyprotein 1ab. Heatmaps represent two groups of serum samples: SARS-CoV-2 infected and control groups. The two left rows of the heatmaps reflect signals obtained with detection antibody only (no serum). Columns represent peptide sequences, rows represent samples. Colours indicate the signal values obtained from triplicate spots ranging from white (0 or low intensity) over yellow (middle intensity) to red (maximal intensity of 65535 light units). The two left rows of the heatmaps reflect signals obtained with detection antibody only (no serum). The upper row in each heatmap reflects the highest possible signal intensity.

**Supplementary Figure 6.**
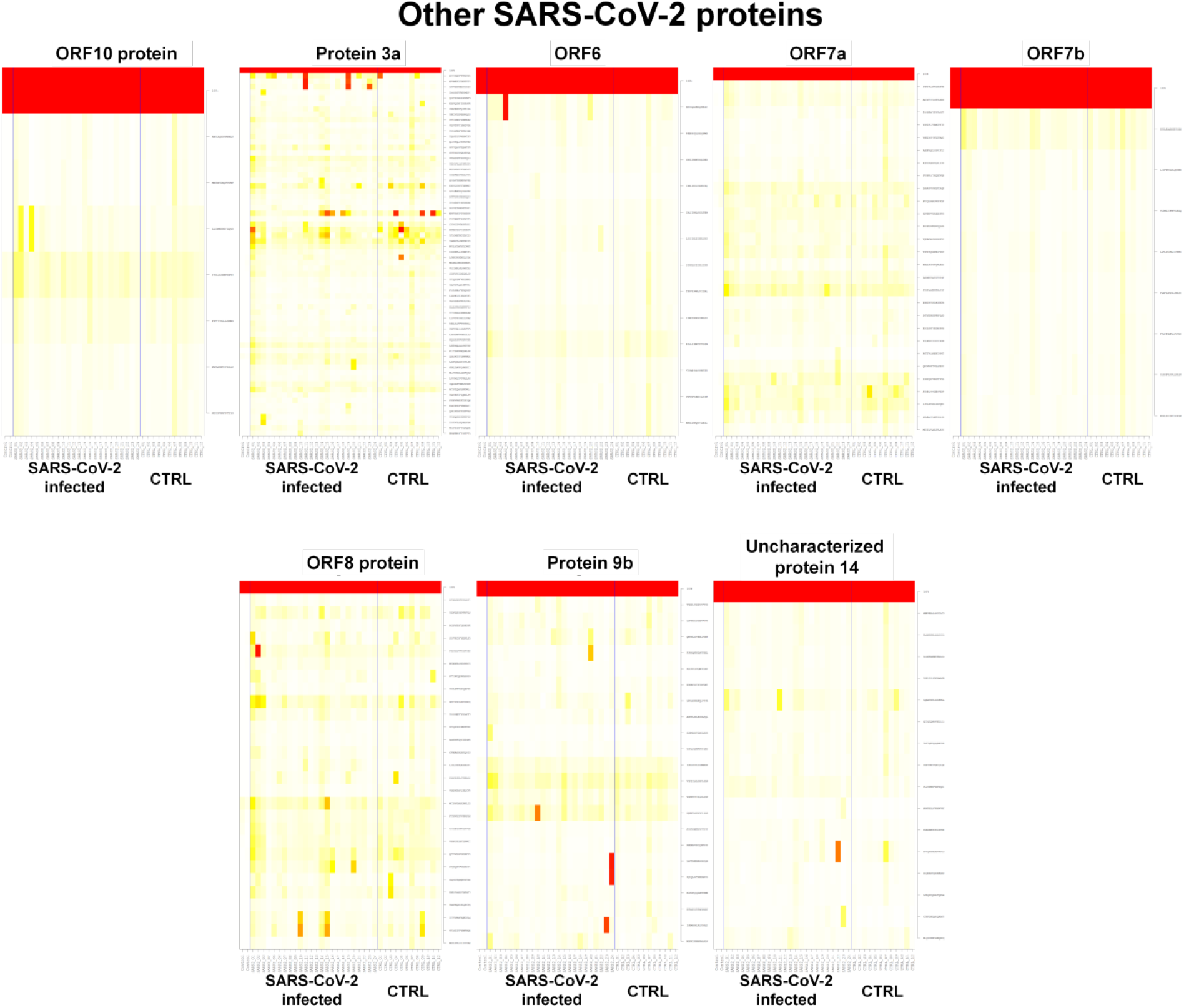
Peptide microarray analysis of IgG responses to other non-structural proteins of SARS-CoV-2. Heatmaps represent two groups of serum samples: SARS-CoV-2 infected and control groups. The two left rows of the heatmaps reflect signals obtained with detection antibody only (no serum). Columns represent peptide sequences, rows represent samples. Colours indicate the signal values obtained from triplicate spots ranging from white (0 or low intensity) over yellow (middle intensity) to red (maximal intensity of 65535 light units). The two left rows of the heatmaps reflect signals obtained with detection antibody only (no serum). The upper row in each heatmap reflects the highest possible signal intensity.

**Supplementary Table 1:** Proteins examined in the study.*

| UniProtKB entry name | Number of peptides | Protein | Organism |
| --- | --- | --- | --- |
| A0A663DJA2_9BETC | 7 | ORF10 protein | SARS-CoV-2 |
| AP3A_WCPV | 66 | Protein 3a | SARS-CoV-2 |
| NCAP_WCPV | 102 | Nucleoprotein | SARS-CoV-2 |
| NS6_WCPV | 13 | Non-structural protein 6 | SARS-CoV-2 |
| NS7A_WCPV | 28 | Protein 7a | SARS-CoV-2 |
| NS7B_WCPV | 8 | Protein non-structural 7b | SARS-CoV-2 |
| NS8_WCPV | 28 | Non-structural protein 8 | SARS-CoV-2 |
| ORF9B_WCPV | 22 | Protein 9b | SARS-CoV-2 |
| R1A_WCPV | 1099 | Replicase polyprotein 1a | SARS-CoV-2 |
| R1AB_WCPV | 1772 | Replicase polyprotein 1ab | SARS-CoV-2 |
| SPIKE_WCPV | 316 | Spike glycoprotein | SARS-CoV-2 |
| VEMP_WCPV | 16 | Envelope small membrane protein | SARS-CoV-2 |
| VME1_WCPV | 53 | Membrane protein | SARS-CoV-2 |
| Y14_WCPV | 16 | Uncharacterized protein 14 | SARS-CoV-2 |
| NCAP_CVHSA | 103 | Nucleoprotein | SARS-CoV |
| SPIKE_CVHSA | 311 | Spike glycoprotein | SARS-CoV |
| VEMP_CVHSA | 17 | Envelope small membrane protein | SARS-CoV |
| VME1_CVHSA | 53 | Membrane protein | SARS-CoV |
| NCAP_CVHOC | 110 | Nucleoprotein | OC43 |
| SPIKE_CVHOC | 336 | Spike glycoprotein | OC43 |
| VEMP_CVHOC | 19 | Envelope small membrane protein | OC43 |
| VME1_CVHOC | 55 | Membrane protein | OC43 |
| NCAP_CVEMC | 100 | Nucleoprotein | MERS |
| SPIKE_CVEMC | 336 | Spike glycoprotein | MERS |
| VEMP_CVEMC | 18 | Envelope small membrane protein | MERS |
| VME1_CVEMC | 52 | Membrane protein | MERS |
| NCAP_CVH22 | 95 | Nucleoprotein | 229E |
| SPIKE_CVH22 | 291 | Spike glycoprotein | 229E |
| VEMP_CVH22 | 17 | Envelope small membrane protein | 229E |
| VME1_CVH22 | 54 | Membrane protein | 229E |

\*: The mnemonic organism identification code of SARS-CoV-2 has been changed. In the updated version, all UniProtKB entry names of SARS-CoV-2 contain the "\_SARS2" species identification code instead of "\_WCPV".

**Supplementary Table 2: Statistical analysis of all data to identify immunodominant epitopes**. The peptides are ranked according to the calculated accuracy value which equals the ratio of the sum of all true positive and true negative cases to the total number of instances. An accuracy of 0.4 is set as a cut-off value for delimitation of the sufficient immunogenicity. 81 Peptides were identified.

See appended Excel list.

**Supplementary Table 3: Patient demographics**.

See appended Excel list.

